# Inhalation exposure to methamphetamine in contaminated residences

**DOI:** 10.64898/2026.10.02.26364615

**Authors:** J.E Dennison

## Abstract

Current health risk assessments from thirdhand exposure to methamphetamine (meth) to occupants of contaminated residences when no cooking or use is still occurring assume that inhalation exposure is insignificant. After Wright et al. (2021) reported substantial meth concentrations in a contaminated residence, the present study was conducted to further test the hypothesis that concentrations were high enough to contribute to total occupant dose. Air sampling was conducted in contaminated residences that had been previously sampled for surface contamination. Samples were collected on personal sampling pumps in fixed locations on treated glass fiber filters is cassettes and analyzed by gas or liquid chromatography with mass spectroscopy. Sixty-nine samples were collected from eighteen residences for the study. Meth concentrations ranged from 0.02 to 15 μg/m^3^. Concentrations in supply air vents were significantly higher than in return vents or in ambient air. Residences with no HVAC systems (baseboard heat) had airborne concentrations similar to residences with HVAC systems turned off, and lower than when HVAC was on. Whether a residence was occupied or not did not show differences. All data from residences with HVAC had a significant correlation with surface concentrations (R^2^ = 0.61) but some factors that affected results were not determined in the study. In the context of the risk assessment and regulatory standard established by California and adopted in several other states, the inhalation exposure would approximately double total occupant dose for infants, and this population would exceed the California reference dose by approximately 100%. Further studies should be conducted to refine these estimates and determine the factors that influence airborne concentrations.

## INTRODUCTION

U.S. states started mandating remediation of meth-contaminated buildings around 1990 (Washington 1989; Oregon 1989) and typically used the minimum detectable concentration of surface residues as their regulatory contamination standards (Hammon and Griffin, 2007), as no health risk assessment had yet been published. Regulatory standards were thus set at 0.05 – 0.1μg/100 cm^2^, the reporting limits for labs performing the analysis of wipe samples. In 2007, Colorado performed a health risk assessment (Hammon and Griffin 2007) to develop a regulatory standard based on adverse health effect data and promulgated a standard of 0.5 μg/100 cm^2^ (CDPHE 2026). Shortly thereafter, California performed a separate health risk assessment and promulgated a standard of 1.5 μg/100 cm^2^ (CalEPA 2009a, 2009b). In both health risk assessments, occupant exposure was based on dermal and ingestion exposure, and inhalation exposure was taken to be insignificant. This assumption may have been partly due to lack of airborne meth data but also based on the chemical’s physicochemical properties and awareness that contamination seemed to remain in buildings for years, suggesting that it was not sufficiently volatile and airborne to rapidly dissipate by exfiltration.

Meth exists in two related molecular forms; methamphetamine (free base) and methamphetamine hydrochloride. The free base (meth_FB_ CASRN 537-46-2) is a secondary amine (Fig. 1) which is a colorless liquid at room temperature (EUDA 2026). In order to facilitate handling, cutting, storage, and distribution, manufacturers react methamphetamine with hydrochloric acid to produce the chloride salt (meth_HCl_ CASRN 300-42-5), a crystalline powder or bulk material (Fig 2). Current analytical methods used for most meth analysis report the meth concentration of both forms combined and do not differentiate between meth_FB_ and meth_HCl_ or any of its optical isomers. Herein, “meth” refers to all forms. Thus, the proportions of meth_FB_ versus meth_HCl_ in air and surface contamination have not been determined.

**Figure 1.**
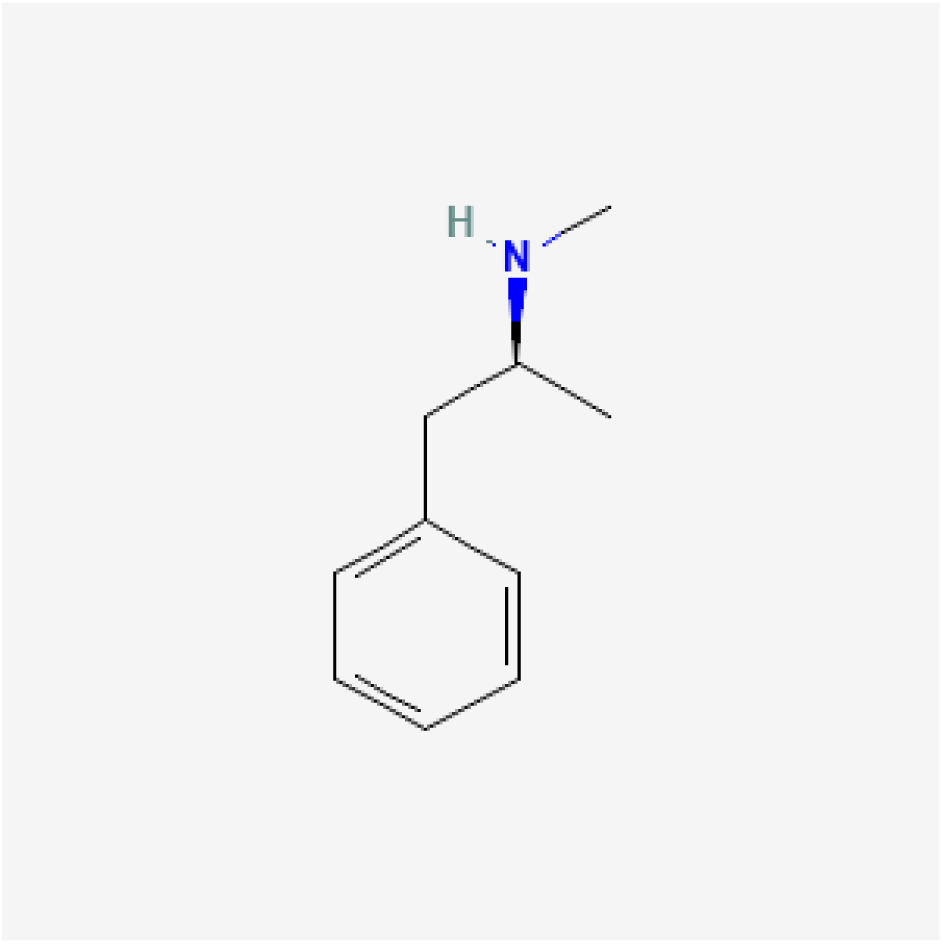
Molecular structure of methamphetamine_FB_. PubChem 2026.

**Figure 2:**
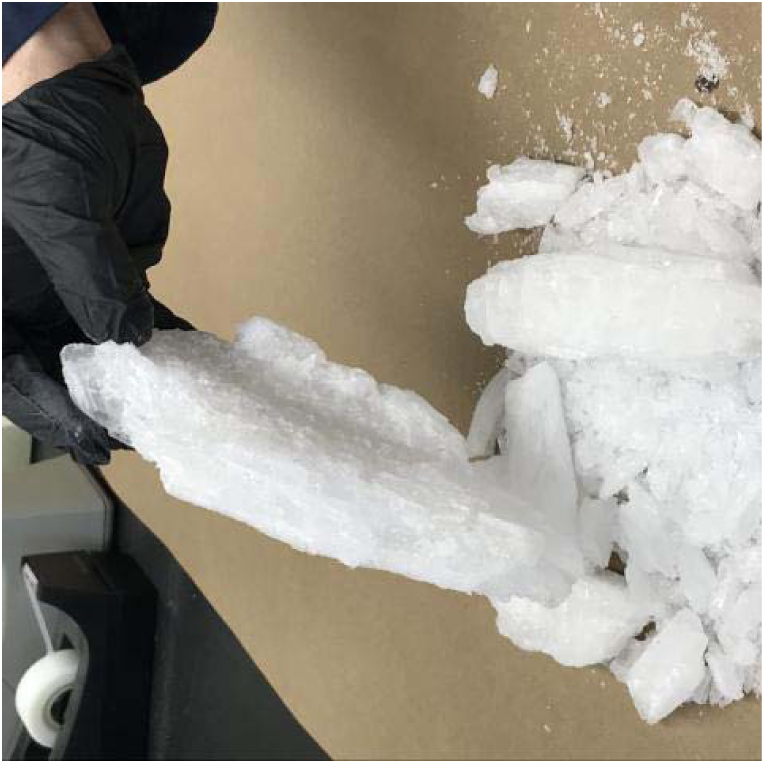
Pure crystal meth_HCL_. USDEA 2026.

Little data are currently available concerning airborne meth in contaminated residences. Several studies have assessed air concentrations after laboratory simulations of manufacturing or smoking of meth, but these have been short term (up to 24 hours) studies where equilibrium conditions in typical residences were not investigated (Bitter, 2017; Martyny, 2008; Van Dyke 2008). Beaudreau et al. (2025) investigated airborne meth concentrations in public transit buses and trains. Wright et al. (2021) were the first to report in a peer-reviewed journal detectable airborne meth from a contaminated residence. The residence had an average surface contaminant concentration of 12.6 μg/100 cm^2^ and air concentrations ranging from 0.5 to 8.3 μg/m^3^. The authors noted that the air concentrations would result in occupant inhalation dose that exceeds the amount recommended by the California health risk assessment. However, on its own, the surface contamination in that residence would also result in excess exposure, as the contaminant level was ∼10-fold higher than the California regulatory standard. Recently, Nam et al. (2026) reported airborne meth concentrations in contaminated residences in Australia, including two cookhouses (where meth was reportedly manufactured) and several smokehouses (where meth was suspected of only being smoked). Their results indicated much higher ratios of airborne meth to surface contamination in cookhouses than smokehouses. The smokehouses had significant inter-house and intra-house variability for unknown reasons. Active ventilation was not in effect in the residences tested.

This study extends the previous work by sampling residences under differing ventilation conditions, by sampling occupied residences where they existed, and by sampling supply air and return air in residences with forced air ventilation or ambient air in unventilated residences. The intent was to explore initial explanatory hypotheses on factors that may influence airborne concentrations as well as to explore the correlation between surface and airborne meth concentrations.

## METHODS

Air sampling was conducted in Colorado residences consisting of single-family homes, condominiums, and rental apartments that had previously been reported to be contaminated. Colorado properties that have had meth activities found (smoking, cooking and related activity) are required by Colorado Department of Public Health and Environment (CDPHE) regulation to be tested via a Preliminary Assessment (PA) (CDPHE 2026). A PA requires wipe sampling in each room in the residence, as well as in HVAC systems, attics, crawlspaces, garages etc. if present. A composite wipe sample is collected in each room by wiping 100 cm^2^ of surface in four locations and combining the wipes in one container. If the sample result exceeds the regulatory standard, that room is non-compliant and must be remediated and shown to become compliant through testing prior to building reoccupancy. PA reports were downloaded from the CDPHE online repository (CDPHE 2025). Each residence’s average surface contaminant concentration was determined by averaging the sample data from all rooms in the residence served by the heating system, which were either forced hot and/or cold air, or baseboard heat or wall heating/cooling units. Rooms that are not on the system such as garages were not included in the average. If any aliquot in a PA’s composite sample was collected in a hotspot, such as inside the bathroom exhaust fan, that sample was not included in the average because hotspots can substantially bias the result (Dennison et al. 2025). The average contaminant concentrations in the residences in this study ranged from 0.2 to 25 μg/100 cm^2^. All residences were tested with owner permission.

Air samples were collected on glass fiber filters (SKC 225-9004) treated with sulfuric acid to stabilize meth on the filter in 37-mm polystyrene cassettes, connected to a sampling pump (Gilian BDX2) with Tygon^TM^ tubing, and calibrated with a primary calibrator (TSI 4140). The flow rate was ∼2 LPM and the sample duration was based on the surface concentrations to keep the estimated analyte mass in the laboratories’ detectable range. Samples were analyzed either by ALS Environmental (Salt Lake City, UT) by LC/MS, or Bureau Veritas (Novo, MI) by LC/MS/MS, to obtain a lower limit of detection. The limit of detection was 0.1 ug and 0.001 μg/sample for the two labs, respectively. Field blanks were reported to contain no detectable meth. Only two samples from contaminated residences contained no detectable meth, and the limit of detection was used for data analysis on these samples. Samples from one residence were omitted from data analysis after review of the PA report showed that biased sampling had been performed by sampling frequently cleaned surfaces (laminate floors and counters). Samples of supply or return air were collected in vents just inside the duct face with the vent cover removed. In a small number of cases when the furnace was not running or the residence had baseboard heat, ambient samples were collected in the center of a room, 3’ from the floor. As ambient air is functionally equivalent to return air, any ambient samples were grouped with return air data. Overall, 69 total samples were collected on 22 days in 18 different residences.

The following comparative sets of samples were collected:

1. Ventilation system on/off/on auto. In all residences combined, 51 samples were collected in supply or return/ambient air in eight residences with the furnace running continuously (“on”) or on automatic (“auto”) which cycles the systems in accordance with heating or cooling demand, and ten samples were collected in four residences with the furnace off.
2. Baseboard heat. Six residences with baseboard heat were sampled (two ambient samples per unit).
3. Duplicates. The reproducibility of collocated samples was used to characterize Sampling and Analytical Error (SAE) with eight sets of collocated samples.
4. Supply versus return air. Thirty-four samples from return vents and 27 samples from supply vents with the HVAC operational were collected.
5. Different supply or return ducts in the same residence. Fifteen pairs of samples were collected in different ducts at the same time.
6. Day-to-day variability. This was investigated in one residence that remained unmitigated for a period of time. The HVAC system was operated in three ways: off (1 day), on (2 days), and on auto (2 days), and 4-6 samples were collected in supply and return vents on each date.
7. Differences between treated and untreated glass fiber filters. Duplicate samples in one return and two supply vents were collected in one residence.
8. Occupied versus non-occupied condition. Two residences were available for testing while occupied. Two to three samples were collected in each of these residences.

Data evaluation was based on either air concentrations (μg/m^3^) or the air concentration to surface concentration ratio (ASR). The units for ASR are μg/m^3^/μg/100 cm^2^, which simplifies to 10^-4^cm^-1^. ASRs are proportional to air concentrations because the denominator of the ratio is fixed for each residence, so any differences calculated are equivalent in either unit of measure.

Data were lognormally distributed so geometric means and geometric standard deviations are reported. Results of any statistical tests that were performed are described; otherwise, differences in results are considered qualitative or nominal, and not tested for significance due to the small dataset.

Risk calculations were based on USEPA inhalation dose calculation methodology (USEPA 1992). Infant inhalation rates and body weights were obtained from USEPA guidance (USEPA 2008). Exposure was assumed to be 24 hours/day. Colorado and California Reference Doses and oral/dermal dose calculations were obtained from published risk assessments (Hammon and Griffin 2007; CalEPA 2009a).

## RESULTS

Data for residences with forced air ventilation are summarized in Table 1 for supply and return vents under the three different operating conditions performed (off, on continuously, and on auto). Geometric means (GMs) of ASRs for supply ducts ranged from 0.30 to 0.72/10^4^cm and were ∼2-fold higher than for return ducts, which ranged from 0.15 to 0.43/10^4^cm under both applicable operating conditions. Supply and return data were statistically different (Welch test p= 0.003 confirmed by Mann Whitney). When the HVAC was operating in auto mode, average results for all vent types were 2.6-fold higher than when the system was turned off and 2.6-fold lower than when the system was operating continuously.

**Table 1:** ASRs (GM) for various HVAC operating conditions in supply and return or ambient locations

|  | # samples | GM | GSD |
| --- | --- | --- | --- |
| **HVAC Auto** |  |  |  |
| Supply vents | 18 | 0.30 | 2.3 |
| Return vents | 18 | 0.15 | 1.7 |
| All vents | 36 | 0.21 | 2.2 |
| **HVAC On** |  |  |  |
| Supply vents | 7 | 0.72 | 2.6 |
| Return vents | 8 | 0.43 | 2.6 |
| All vents | 15 | 0.55 | 2.6 |
| **HVAC Off** |  |  |  |
| Return vents | 10 | 0.081 | 6.2 |
| **All data** |  |  |  |
| Supply vents | 27 | 0.37 | 2.5 |
| Return vents | 34 | 0.16 | 3.6 |
| All vents | 61 | 0.23 | 3.3 |

In Residence 1, return duct ASR with the furnace on auto on two days averaged 0.19/10^4^cm whereas the return duct ASR on two days with the furnace on continuously averaged 0.31/10^4^cm. While the constantly running furnace yielded results that are nominally higher than when the furnace cycles according to temperature demand, the difference is confounded by day-to-day variability.

The GM of ASR for baseboard heated units was 0.18/10^4^cm. When baseboard units were combined with HVAC units, the overall GM of ASR was 0.21/10^4^cm (GSD 3.2). When baseboard units were combined with only return data, the overall GM of ASR was 0.15/10^4^cm (GSD 2.0).

Eight sets of duplicate samples were collected at the same time in the same locations (side-by-side) as a measure of SAE. The relative percent differences of ASRs ranged from 0-17% with an average of 8.1%. Fifteen sets of samples were collected in ducts (supply or return) in the same residence at the same time. Samples in supply were compared to other supply samples and return samples with other return samples. Paired t-test found no significant difference between samples in different ducts of the same type (p=0.63). The relative percent difference between ASRs obtained from treated and untreated GF filters ranged from 10-18%. This is similar to the overall SAE. Nominally, the untreated filters gave slightly higher results than the treated filters.

Somewhat inconsistent results were obtained for samples from different days in Residence 1 (Fig. 3). With the same HVAC operating conditions, average results varied by ∼2-fold. Within days, concentrations in samples in supply ducts exhibited relatively low variance and samples in return ducts also exhibited low variance, as for the overall dataset. The ASR GM for occupied residences was 0.19/10^4^cm (GSD 1.9).

**Figure 3:**
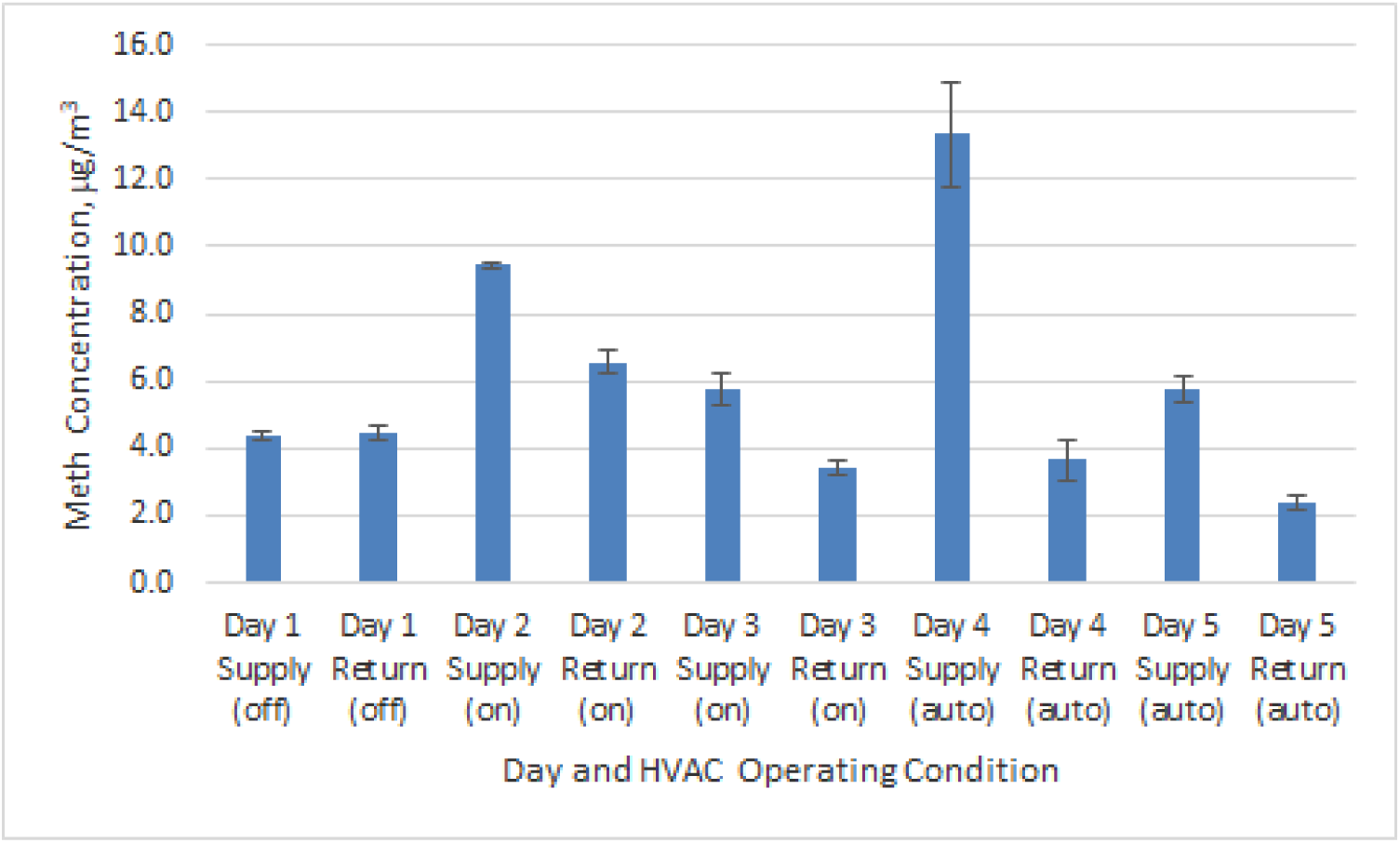
Meth air concentrations (average, ug/m3) on different days and HVAC conditions in Residence 1

Data from residences on HVAC systems were examined for correlation (Fig. 4). As ambient background meth air concentrations in uncontaminated residences are <1 μg/m^3^, the regression equation was forced through zero and only one day’s tests from Residence 1 were included to avoid bias. The regression equation was Air Concentration (μg/m^3^) = 0.17*Surface Concentration (μg/100 cm^2^). R^2^ was 0.61. The correlation was statistically significant (p=1^10^-9^).

**Figure 4:**
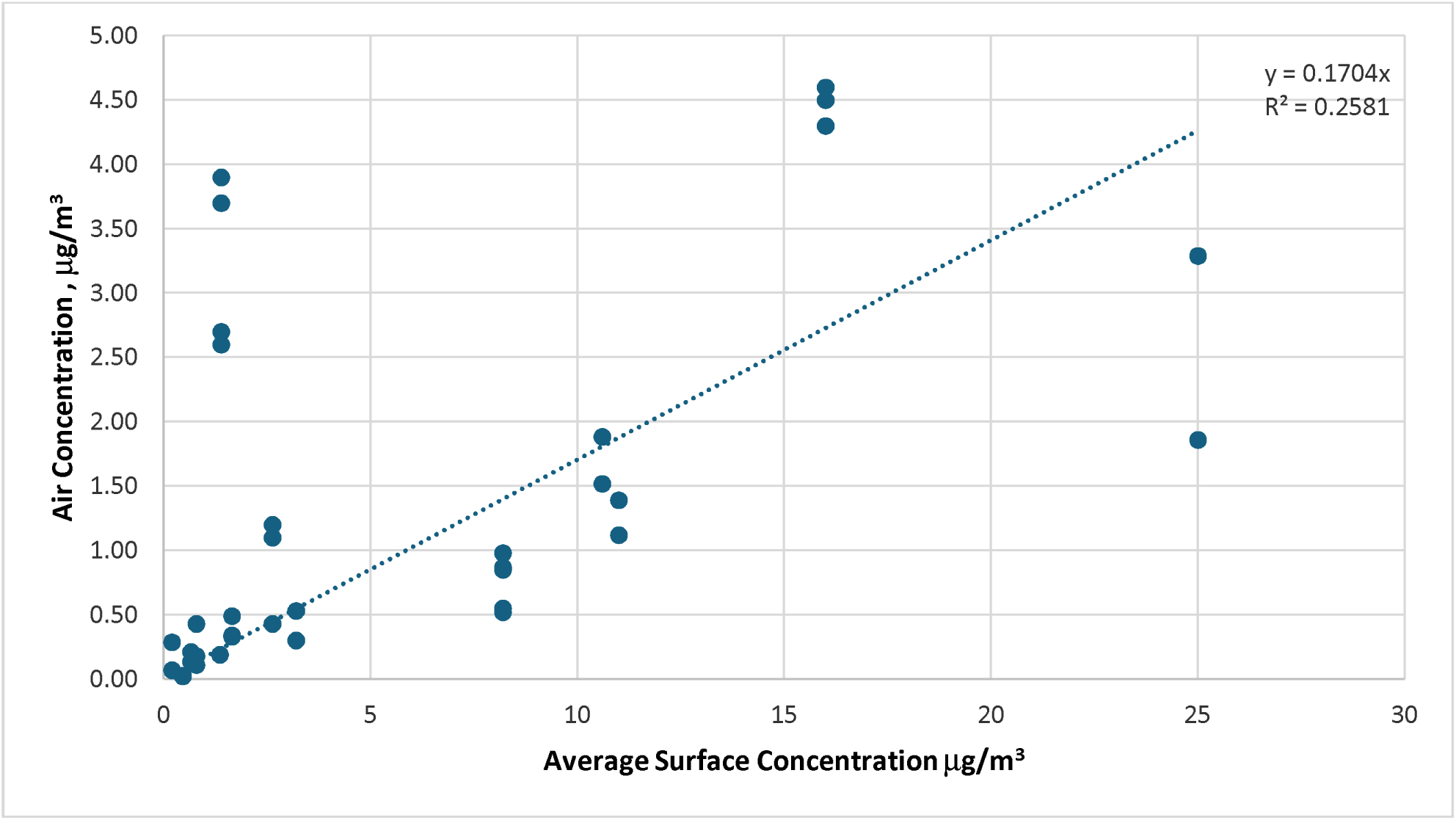
Correlation between airborne meth and surface concentrations for all residences on HVAC

By inspection, if data were restricted to residences with surface concentrations <3 μg/100 cm^2^, the ASR would be higher (data not shown). Insufficient data were available in this study to evaluate non-linear models.

Table 2 provides the results of incremental dose calculations based on the ASR from the linear regression (0.17/10^4^cm) at regulatory standards for Colorado and California, for infants. The total dose, when inhalation was not included, calculated by Colorado and California health departments is listed as “Oral and Dermal Dose” (the latter using the California 95^th^ percentile estimate). Based on the existing risk assessments, inhalation is estimated to add 44% and 95% additional dose for Colorado and California, respectively. However, the target dose (Reference Dose (RfD)) allowed by the two health standards vary widely, which Colorado targeting 5-70 μg/kg/day and California targeting 0.3 μg/kg/day. Therefore, the estimated total dose including inhalation in relation to Colorado (0.29 μg/kg/day) is well below the Colorado RfD whereas the California dose exceeds the California RfD. This is caused partly by the differences in RfDs but also because more inhalation dose in received at the higher California regulatory standard.

**Table 2:** Incremental inhalation dose calculations at Colorado and California regulatiory standards

| Parameter | Units | Colorado | California |
| --- | --- | --- | --- |
| Air/Surface Ratio (ASR) | /10<sup>4</sup>cm | 0.17 | 0.17 |
| Surface Concentration | µg/100 cm<sup>2</sup> | 0.5 | 1.5 |
| Air Concentration, calculated | µg/m<sup>3</sup> | 0.085 | 0.255 |
| Body Weight | kg | 6.75 | 6.75 |
| Inhalation Volume | m<sup>3</sup>/day | 7.0 | 7.0 |
| Inhalation Dose | µg/kg/day | 0.088 | 0.264 |
| Oral and Dermal Dose | µg/kg/day | 0.2 | 0.278 |
| Total Dose | µg/kg/day | 0.29 | 0.58 |
| Increase in total dose | % | 44% | 95% |
| Reference Dose | µg/kg/day | 5-70 | 0.3 |

## DISCUSSION

Beaudreau et al. (2025) investigated airborne and surface meth contamination on trains and buses. Lower meth surface (0.02-0.13 μg/100 cm^2^) and air contamination (0.03-0.24 μg/m^3^) was found than in this study, possibly due to higher effective ventilation rates in public transport than in residences, different cleaning practices, or other reasons. The combined ASR was 1.8/10^4^cm and is higher than in this study, possibly due to the lower surface concentrations in that study or possible meth use during testing.

Wright et al. (2023) reported an average air concentration of 4.7 μg/m^3^ in a residence that had an average surface concentration of 12.6 μg/100 cm^2^ (ASR = 0.37/10^4^cm). The most comparable data in the present study would be the return average ASR of 0.16/10^4^cm, but the result in Wright’s study matched the GM of supply vents in the present study (0.37/10^4^cm).

There are many reasons a direct comparison is not possible between the two datasets however. The Wright et al. (2023) study used XAD2 sorbent in an ORBO 49P SupelcoPak-20 tube as opposed to treated glass fiber filters. The method of averaging surface concentrations was not reported, and surface sampling locations may not be comparable. Moreover, the cited study’s dataset was small, consisting of five samples in one residence. However, it is interesting to note the similarity of the results despite methodologic differences. The Wright et al. data did not yield a consistent pattern for airborne meth relative to HVAC system operation, ranging over more than an order of magnitude.

Nam et al. (2026) reported a study of meth in air in contaminated residences in Australia. Forty-two air samples were collected in an unspecified number of residences, mostly 1-2 samples per residence, in two subsets (smokehouses and cookhouses.) The ASR for smokehouses was 0.018/10^4^cm (R^2^=0.13, presumably forced through 0). For cookhouses, the ASR was dramatically higher at 1.08/10^4^cm (R^2^ = 0.80 presumably forced through 0). The reason for the high cookhouse ASR is not simply that cookhouses often have much higher surface concentrations than smokehouses have. They do, but this is factored out by the ASR ratio. It need be noted that the cookhouse data consisted of two samples in each of two residences, with high surface concentrations (18-60 μg/100 cm^2^). Whether ASR for cookhouses would remain high when surface concentrations are close to the regulatory standard is unknown. It is worth noting that the residences in the Nam study did not have “active ventilation,” but some windows were opened and the impact of this operating condition is unknown.

One confounder of differentiating cookhouse and smokehouse results is not having definitive knowledge of whether cooking occurred or not. The author of the present study has encountered a few cases where police reports stated that cooking occurred, and the residences had no actual evidence of manufacturing or detectable meth. The author also assessed a residence where cooking very likely occurred, for a short period of time, that was not contaminated above the regulatory standard. Conversely, when it is reported that only smoking occurred, the information is often provided by someone who was not present, and may have motivations to obscure information. It stands to reason that where meth is cooked, it is also smoked in nearly all instances, but the relative frequency of smoking and cooking is unknown. Thus, anecdotal information is not always reliable and/or complete.

There are several methods for cooking meth reported by drug enforcement agencies, including the hydriodic acid/red phosphorus, iodine/red phosphorus, iodine/hypophosphorous acid, birch, phenyl-2-propanone, (NDIC 2005) and the one-pot methods (USDEA 2020). Van Dyke et al. (2009) reported emissions from cooking with the red phosphorus method, but currently, the one-pot (also known as “shake and bake”) method is used most often (Recovery Village) and no data are presently available on surface contamination from methods other than the red phosphorous. Thus, cookhouses may not have had cooking and smokehouses could have been cookhouses at times, so the distinction is problematical. In the residences included in this study, no cooking was reported and based on the scarcity of law enforcement apprehension of meth labs since 2006 (Dennison 2025), most were likely smokehouses.

A second confounder of study comparison is sampling methodology. Nam et al. (2026) used OVS XAD-2 Tubes, containing a quartz filter and XAD sorbent. Wright et al. (2023) used ORBO-49P (OVS) Supelpak tubes with a glass filter and XAD2 sorbent. Beaudreau et al. (2025) used nylon filters. Raynor and Carmody (2006) used silica gel followed by glass fiber filters.

This study used glass fiber filters coated with sulfuric acid, similar to Van Dyke et al. (2009). The intent of the acid treatment is to stabilize any meth_FB_ that may be present by conversion to methamphetamine sulfate. Jeronimo et al. (2024) reported method validation studies performed for air sampling meth and several other opioids on glass fiber and other media. Their studies found that 100% collection efficiency was attained after passing clean air through glass fiber filters spiked with meth, as well as 94% extraction efficiency, and reported that their method met OSHA method validation criteria (OSHA 2010). Additional studies have been performed with glass filter and tube sample trains that found <1% of the total analyte detected was detected on the tube and 99% was detected on the filter. (Jeronimo, 2026). While analytical methods were not the same in all studies, they all relied on gas or liquid chromatography/mass spectroscopy and may not be responsible for much of the differences found between studies. It appears that use of treated filters may not have an impact, as treated and untreated filter yielded similar results.

The issue of airborne contaminant capture is slightly complicated since meth is present in two forms. Meth_FB_ is a volatile organic compound (VOC) but with relatively low vapor pressure. Both stereochemical forms, *d* and *l* optical isomers, and racemic mixtures for both meth_FB_ and meth_HC_.are present. However, methods of wipe sample analysis do not differentiate between any of the forms and all forms are detected and reported as meth. What form(s) of meth are present in contaminated buildings has not been investigated, but meth_HCL_ is inevitably present since this is the form that is smoked and cooked. How much meth_HCL_ may be transformed into meth_FB_ is unknown. Meth_HCL_ would be associated with all particulate matter (PM) in the property, which may be airborne PM or adhered to surfaces. Meth_FB_ may be airborne as a vapor or adsorbed/absorbed to PM or surfaces.

The lack of information on what forms of meth are present and the concentrations associated with PM in air or in settled dust creates challenges in understanding what building-related factors control the overall airborne concentrations. One might suspect that air concentrations in supply air may be lower than in ambient and return air because the supply air has passed through HVAC filters, removing some PM. The removal efficiency will depend in part on particle size and which form of meth is present. Surprisingly, the current study indicated that air concentrations in supply air were higher than in ambient and return air. This is correlative to the higher concentrations found when the HVAC system was on auto versus off and on continuous versus auto. When the system is not supplying air (i.e., the fan is off), a “supply” vent sample is actually sampling ambient air after room air and vent air are mixed, so the more the fan operates, the more actual supply air is sampled. The reasons for the higher concentrations in supply air are unclear but could be related to the concept of an equilibrium between concentrations in the HVAC system compared to the rest of the residence. It has been previously reported that meth concentrations in HVAC ducts averaged 10-fold higher than the average for the ordinary surfaces in the same property (Dennison 2026). However, the rate and therefore length of time it takes for meth to build up in the system when meth activity is occurring is unknown. After meth use ceases and air exchange occurs between the residence and outside the residence, indoor ambient concentrations would decline, and the direction of flux could shift from net influx to the HVAC to net outflux. If this mechanism is significant, timing of air sampling via a vis meth activity could influence the accuracy of air data to represent long term exposure.

The unknown accuracy of wipe sampling data also hinders understanding results of this study. The PAs that provided the wipe sample data followed state sampling requirements wherein each room is sampled in four locations, arbitrarily selected by the sampling professional. Review of PA reports indicated that sample locations generally did not include hotspots with high concentrations of meth. However, comprehensive studies of the ability of 4-part composite wipe sample data to estimate the true average surface concentration have not been conducted and the variance is unknown. Low bias in some reports could be responsible for some of the atypical results. Unequal room sizes also introduce a bias when the arithmetic mean of room surface concentrations is used to estimate the whole residence average.

A statistically significant correlation was found between air and surface concentrations in this study. Conversely, in Residence 1 that was sampled five times, day-to-day differences were surprisingly large, given the low SAE and comparability of sampling in different vents on the same day. Lack of occupancy did not appear to have an effect, although additional data are needed. Additional variables that may explain the day-to-day variation were not investigated in this study but may include 1) activity that affected entrainment of PM of which the investigator was not aware 2) outdoor temperature (and possibly humidity) variation that affects HVAC run conditions and functionality. Atypical high results were obtained in four residences (Fig. 4).

These may be caused by errors in surface concentrations estimates, in air sampling data, or unknown factors or may be correct. Unfortunately, in this study confirmation sampling was generally not possible as owners typically initiated remediation before sample results were available.

As previously mentioned, there is not a clear rationale for the large difference between Nam et al. estimates and this study’s. However, this study agreed better with Beaudreau et al. (2025) and Wright et al. (2023). The Nam study had very low correlation in data from smokehouses, whereas this study had significant correlation. It can be noted as well that while a method can under-sample an airborne contaminant, it cannot over-sample and the similarity in ASR results based on data obtained from two independent labs in this study provides additional confidence.

Wright et al. (2023) noted that at elevated contaminant levels, children would be expected to receive 20% of the total meth dose by inhalation, and adults ∼60%. While inhalation doses indicated by the present study are not a concern in relation to the Colorado risk assessment, they are a clear concern in relation to California’s assessment, resulting in exposure above the Reference Dose. It is just as much of a concern that the two risk assessments suggest acceptable exposures for infants or young children that are ∼20-200 fold different. Which methodology is the most appropriate between the two risk assessments is outside the scope of this study. Given the potential for exposures exceeding the California RfD, additional studies concerning airborne meth, especially in residences that are contaminated at concentrations near the regulatory standard need to be performed. Future studies may be able to unravel the forms of meth present (meth_FB_ , meth_HCL_, and particulate bound meth), the factors that enter into day-to-day variations, and the relationship between HVAC concentrations and ambient concentrations over time.

This study suggested that there is some correlation found between air and surface concentrations at all levels of contamination. If variables that affect the air concentrations are determined and controlled, an improved correlation should be obtainable. As noted, the variance in wipe sample data is unknown, so at this point, how much missing correlation is caused by the wipe data is unclear. The potential exists that air sample data could actually be more reliable as an indication of average contaminant levels than wipe sample, since air concentrations are much more homogeneous than surface concentrations are. If air sampling under the proper operating conditions was found to exhibit less variance than wipe sampling, meth testing might become less costly with much less time required and fewer samples needed to adequately characterize the contamination status of a property.

## LIMITATIONS

Several limitations were encountered in this study. The use of different air sampling methods impacts comparability to other studies. The lack of knowledge on the form(s) of meth present in the residences, and their relative association with PM affects both hypothesis generation as well as interpretation of results. Lack of knowledge regarding the factors that affect airborne meth concentrations also leave it uncertain what conditions would represent average air concentrations for future occupants experiencing thirdhand exposures. The variance associated with PA wipe sampling being unknown offers a potential of bias. Additional data are needed, especially at the surface concentrations of regulatory standards as well as in occupied contaminated residences.

## RECOMMENDATIONS

Recommendations that grow out of this study include: 1) development of air sampling methods that can discriminate between meth_FB_, meth_HCL_, and both forms associated with PM; 2) assessment of the variance in meth wipe sampling; 3) harmonization of the health risk assessments and consideration of the inhalation component of total dose; 4) additional studies to develop insight into the factors that influence the airborne concentrations; 5) additional studies to strengthen confidence in the correlation between air and surface concentrations.

## CONCLUSIONS

There is an apparent disparity between the results of Nam et al. (2026) and this study, with the present result expressed as ASRs much higher than the previously-reported smokehouse ASRs and much lower than the reported results for smokehouses. However, the present results used a better validated sampling methodology, were in the same range as some other studies albeit of limited methodological comparability, and the correlation was statistically significant. Further studies will be needed to improve the estimates.

The present study suggests some factors with HVAC conditions that do not appear to have an effect and others that do. When HVAC was off, air concentrations were lower and concentrations in supply air were higher than ambient and return air when HVAC was on. Occupancy did not appear to have a significant effect. Results of samples on the same day, even in different locations, were quite consistent, but there were substantial day-to-day variations that there is no present explanation for. The data suggest that inhalation exposure may be significant in the context of the California risk assessment and regulatory standard. For this reason, additional studies should be conducted to extend these results.

## Data Availability

All data produced in the present study are available upon reasonable request to the authors

## ACKNOWLEDGMENTS

This work was funded in part by Public Housing Authorities in Colorado. Assistance with sampling from Paul Jaeckel and Ashley Brede was appreciated. The author declares it has no competing interests but is involved in consulting activity.

## DATA AVAILABILITY STATEMENT

Supplementary data are available at [REDACTED].

